# Analysis of the Dynamics, Outcome, and Prerequisites of the first German SARS-CoV-2 Superspreading Event

**DOI:** 10.1101/2021.09.01.21262540

**Authors:** Lukas Wessendorf, Enrico Richter, Bianca Schulte, Ricarda Maria Schmithausen, Martin Exner, Nils Lehmann, Martin Coenen, Christine Fuhrmann, Angelika Kellings, Anika Hüsing, Karl-Heinz Jöckel, Hendrik Streeck

**Author notes:** Corresponding author: Hendrik Streeck, Institute of Virology, University Hospital Bonn, Venusberg-Campus 1, 53127 Bonn, Germany.

## Abstract

**Background:** The beginning of the COVID-19 pandemic was shaped by superspreading events including large-scale outbreaks. In Germany the first SARS-CoV-2 outbreak was a superspreading event in a rural area during indoor carnival festivities in February 2020.

**Methods:** 51 days after the event all known participants were asked to give blood samples, pharyngeal swabs and answer a self-administered questionnaire. Metric room coordinates for all tables, seats, and ventilation-points were assessed.

**Findings:** We analyzed infection rates among all 411 participants, and the risk of infection in relation to various factors including age, alcohol consumption, and ventilation system. Overall, 46% (n=186/404) of the participants had been infected. We demonstrate that the spatial distribution of infected participants was associated with proximity to the ventilation system (represented as inverse distance, with Odds Ratio OR 1.39, 95% KI [0.86; 2.25]). Interestingly, the risk of infection was highly associated with age, whereby children (OR: 0.33 [0.267; 0.414]) and young adults (age 18-25) had a lower risk of infection than older participants resulting in an average infection risk increase of 28% per 10 years age difference. Behavioral differences also impacted the risk of infection including time spent outside (OR: 0.55 [0.33; 0.91]) or smoking (OR: 0.32 [0.124; 0.81]).

**Interpretation:** Our findings underline the importance of proper indoor ventilation for events in the future. The lower susceptibility for children and young adults indicates their limited involvement in superspreading events.

**Funding:** The government of North Rhine-Westphalia (Germany) supported the study with 65,000 Euro.

**Research in context:** *Evidence before this study:* The scientific literature was searched for the term “superspreading event AND Covid-19 OR Sars Cov 2” and identified published papers from China, South Korea, Europe, and North America. Most researchers analyzed superspreading events within a health care setting e.g. in hospitals or nursing homes, or described the general impact of superspreading events on the global pandemic. Only a few metanalyses of transmission clusters analyzed party occasions (e.g. a nightclub in Berlin, Germany) as superspreading events. These reports describe less than 100 infections and are very limited due to missing data or reporting biases. Therefore, the ability to draw scientific conclusions is also limited. Additionally, to our knowledge, there are no studies, which investigated individual behavior, the location, and role of children during a superspreading event. The research for the study started April 2020 and was concluded in June 2021.

*Added value of this study:* Our report analyzes the first COVID-19 superspreading event in Germany in detail, which was not only a unique setting but also included children and adults in the same room. We demonstrate that nearly half of the participants were infected with SARS-CoV-2 and that the proximity of the seating to the ventilation system was an important risk factor for infection. The data showed that low physical distance including singing and duration of attendance at this event increased the risk of infection, while regular smoking and spending the break of the event outside lowered the risk of infection. This underlines the benefit of airing to lower the amount of both droplets and aerosols. Furthermore, we found lower infection in children than adults despite being in the same room suggesting differences in infectability in children. Indeed, we observed that an additional 10 years of age is on average associated with 28% increased risk of infection.

*Implications of all the available evidence:* Taken together, the results demonstrate the importance of the ventilation system during superspreading events. In particular children and young adults had a lower risk of infection during the event indicating that they have a limited role during this pandemic. Overall, our data demonstrate in detail age-dependent infectability as well as highlights to understand transmission dynamics in order to improve comprehensive public health preparedness measures.

## Introduction

Severe acute respiratory syndrome coronavirus 2 (SARS-CoV-2) is a highly transmissible and pathogenic RNA virus that emerged in late 2019 and has caused a pandemic threatening human health and public safety worldwide.^1^ While factors shaping the dynamics of a pandemic are multifactorial, virulence and reproductive number are important properties of a virus.^2^ For SARS-CoV-2 there is a substantial over-dispersion of the secondary infection distribution (individual R0) for an individual infected with SARS-CoV-2^2^. An over-dispersed R0 means that most infected people do not transmit (individual R0 = 0) while a minority of infected people are super-spreaders (individual R0 >5). Superspreading has been observed for many infectious pathogens, such as measles or SARS.^3^ During the SARS pandemic in 2003 a superspreading event was defined as one infected person infecting eight others.^4^ For SARS-CoV-2 it has been estimated that 80% of the infections are caused by 10% of infected individuals highlighting the importance of the cluster factor (k).^2^ In Germany an indoor carnival event in the beginning of 2020 is considered as the first major outbreak in a German city and was considered a hotspot during the beginning of the pandemic in Germany.^5^ Other SARS-CoV-2 superspreading events worldwide have been linked to indoor gatherings with close proximity of individuals.^6^ Nevertheless, most of the reported superspreading events had less than 100 cases and the reports are limited by missing data or a reporting bias.^6^

Here, we closely examined the prerequisite of a unique super-spreading event in Germany during the SARS-CoV-2 pandemic, where nearly half of the participants became infected including children. We systematically analyzed infection rate, potential individual, and environmental risk factors for infection as well as the role of the ventilation system.

## Materials and methods

### Study design and sampling

This cross-sectional epidemiological study was conducted 51 days after a carnival celebration in the beginning 2020. Eleven days after the event authorities sent all known participants into quarantine after testing 38 out of 99 individuals PCR-positive. All adults known to have attended the event were invited to participate in the study. About 450 persons attended the event of which 411 participated in the study (**figure 1**, participation rate 91.3%). All study participants provided written informed consent before enrolment. The Ethics Committee of the Medical Faculty of the University of Bonn approved the study (approval number 085/20).

**Fig. 1:**
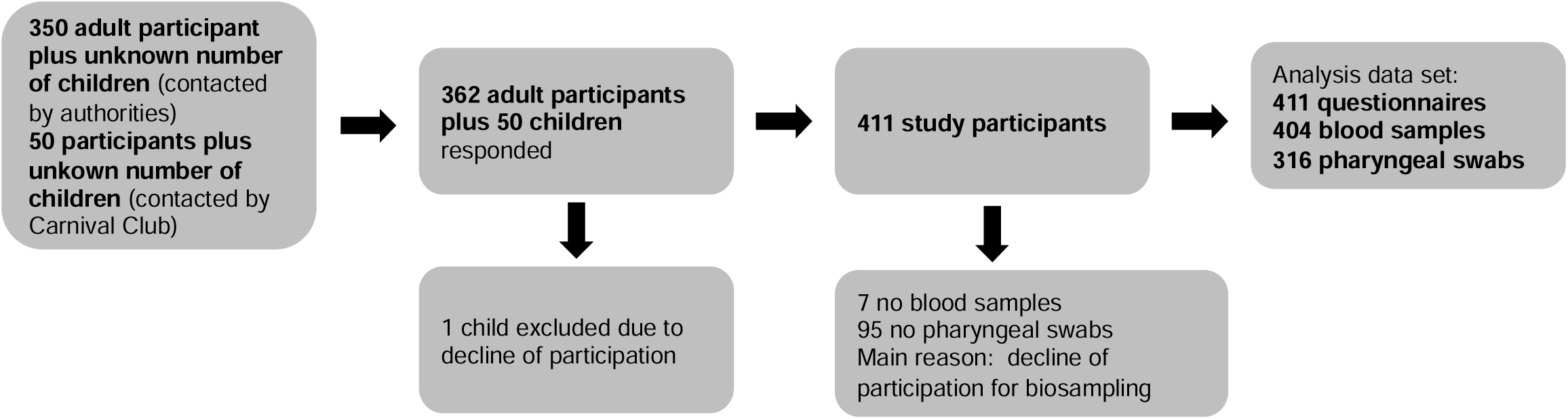
Study participants. Of the 400 people contacted originally (left) 362 adults and 49 children agreed to enroll in the study. An overview of the number of samples collected is given on the right. Downstream sample processing included centrifugation of blood samples for plasma collection (SARS-CoV-2 ELISAs), and viral RNA extraction from swab samples (SARS-CoV-2 RT-PCR).

**Fig. 2:**
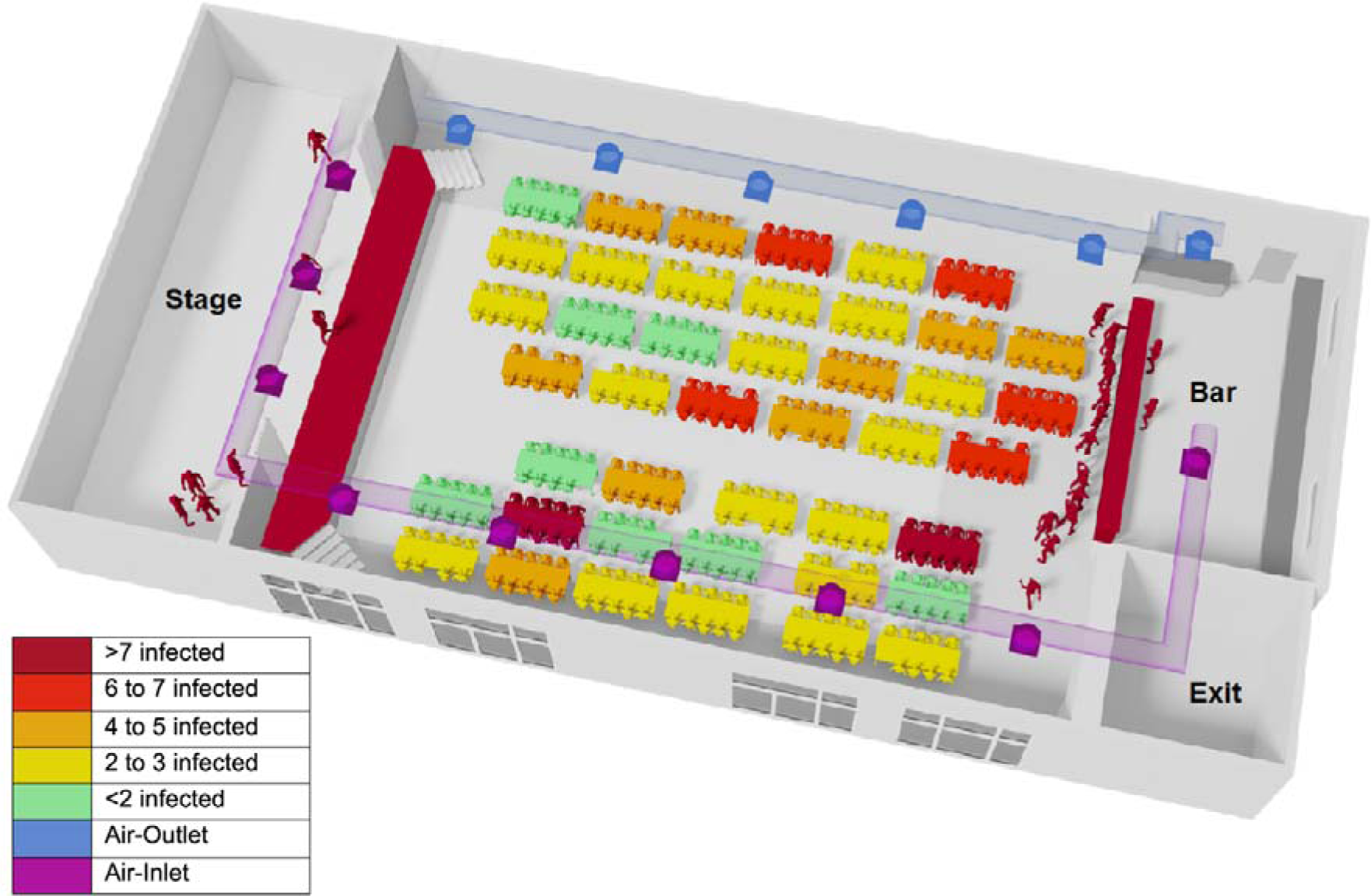
Reconstructed 3D-Model of the venue hall. Self-administered questionnaires included questions about main seating-position of the participants during the evening event as specifying table and seat with the help of a schematic seating plan. Metric room coordinates for all tables, seats, and ventilation-points were assessed and the seating was reconstructed from pictures taken during the event. Therefore, the location of the stage, the bar, the exit as well as the tables and the air-inlets/outlets were reconstructed in a 3D-Model. The original external dimensions of the building were 27m x 13.20m x 4.20m. Tables, where more than 7 infected individuals have stayed are colored in dark red, this includes the stage and bar as well. Air-inlets are colored in violet and the air-outlets in blue. Infected participants had been seated mostly at tables close to the bar, the bar itself and on stage. One table with 8 out of 11 infected people, was located far away from the bar at the other side of the hall and close to an air inlet. The group sitting on stage showed as well high numbers of infection (18 infected out of 24). Greater proximity to air outlets seems to be associated with increased risk of infection with a crude OR=1.39 [0.86; 2.25].

Self-administered questionnaires included questions about demographic background, symptoms of viral infection as well as detailed information about the behavior during the event. Participants’ arrival and exit times were assessed in 1-hour categories. Study participants were asked to provide blood specimens and pharyngeal swabs for further analysis.

### Pharyngeal swab and blood preparation

Pharyngeal swabs of participants were performed with FLOQSwabs (Copan) and immediately stored in UTM RT-mini tubes containing UTM Viral Stabilization Media (Copan) at 4 °C. Venous blood was drawn into EDTA tubes (Sarstedt) per volunteer and was transported to the laboratory at the University Hospital Bonn.

### Anti-SARS-CoV-2 ELISA

Anti-SARS-CoV-2 IgA and IgG were determined using enzyme-linked immunosorbent assays (ELISA) on the EUROIMMUN Analyzer I platform.^5^ According to the manufacturer’s instructions a result was considered positive when a ratio (extinction of sample/extinction of calibrator) of 0.8 or higher was reached. The guidelines of the German Medical Association (RiliBÄK) were abided by, including internal and external quality controls.

### Reverse transcription polymerase chain reaction (RT-PCR)

Viral RNA was extracted from each 300μl swab sample via the chemagic Viral 300 assay (according to manufacturer’s instructions) on the Perkin Elmer chemagic™ Prime™ instrument platform. The presence of two viral target genes (E and RdRP) was assessed in each sample by real time RT-PCR (SuperScript™III One-Step RT-PCR System with Platinum™ TaqDNA Polymerase, Thermo Fisher). The following primers were used, for E gene: E_Sarbeco_F1 and R, and probe E_Sarbeco_P1, for RdRP gene: RdRP_SARSr_F, and R, and probe RdRP_SARSr-P2.^7^ In addition, an internal control for RNA extraction, reverse transcription, and amplification was applied to each sample (innuDETECT Internal Control RNA Assay, Analytik Jena #845-ID-0007100). If amplification occurred in both virus-specific reactions samples were considered positive.

### SARS-CoV-2 Neutralization Assay

A plaque reduction neutralization test was used to determine SARS-CoV-2 neutralization capacity as previously described.^5^ Briefly, plasma samples were heat-inactivated and supernatant transferred to a new tube and serially two-fold diluted in OptiPROTMSFM (Gibco) performed. 120 mL of each plasma dilution was mixed with 80 plaque-forming units (PFU) of SARS-CoV-2 in 120 mL OptiPRO SFM (GIBCO) cell culture medium and seeded with 1.25×10^5^ Vero E6 cells/well. Subsequently, the inoculum was removed and cells were overlayed with a mixture of carboxymethylcellulose (Sigma) and 2xMEM (Biochrom). Following 3-day incubation, the overlay was removed and the 24 well plates were fixed using a 6% formaldehyde solution and stained with 1% crystal violet in 20% ethanol revealing the formation of plaques. Finally, the neutralizing titers were calculated as the reciprocal of serum dilutions resulting in neutralization of 50% input virus (NT50), read out as reduction in the number of plaques.

### Data management and quality control

The Clinical Study Core Unit of the Study Center Bonn (SZB) supported the study by outlining the study protocol and developing the informed consent form as well as participants information sheets with respect to data management and quality control. The data were gathered on paper-based Case Report Forms (pCRF). Data was entered as double-data-entry into the REDCAp study database programmed and hosted by SZB. Study personnel was trained by experienced members of the SZB. A quality manager was on site to support the study team. Monitoring of trial data and informed consent forms was performed according to the monitoring plan by qualified SZB staff. The ethics committee of the Medical Faculty of the University of Bonn was involved and approved the study (reference no. 085/20)

### Spatial information

Metric room coordinates (length and width [m]) for areas, tables, seats and ventilation shafts were assessed via measurements, seating plan and photos from the event. Persons providing multiple positions were considered as spending an equal amount of time on different positions. When exact seating was unclear and information was available on table or greater area localisation (bar, stage), average coordinate values were used.

On the grounds of these coordinates, we calculated pairwise metric distances between all persons and distances to closest inletting and purging airshafts. For all persons their pairwise inverse distances were summarized as mean inverse distance. Inverse metric distances to persons or airshafts were regarded as representing infectious potential through local proximity, and inverse distances were capped at 2.5 (the inverse of the width of a seat of 0.4 m). Alternatively, we counted all, and all infected persons within adjacent rings of 1.5 m width around each participant as a measure of crowdedness and infectious potential.

### Statistical analysis

Associations between positive infection status and exposure variables were analysed via logistic regression models. Exposure variables were included crudely, and adjusted for potential confounding factors age, sex, and duration of attendance as fixed effects. To correct for common household effects a random effects model was used. We present odds ratios with 95% confidence intervals. Because we present data on a single specific event among a limited number of participants, we completely refrain from presenting p-values. All analyses were done with SAS 9.4.

## Results

411 out of estimated 450 participants of the event responded to our study invitation, resulting in a response rate of 91.3%. 404 individuals provided plasma samples and 316 pharyngeal swabs (**figure 1**). Genders were represented equally among all 404 participants (48% were male) with a broad range in age ((range 6-79) median age 36 years) and level of education (**table 1**). 297 individuals were residents of the community the event took place in, 103 lived in other parts of the county, and 11 were external visitors.

**Table 1:** Distribution of demographic factors and exposure information of interest among study participants who tested positive or negative in serology test of SARS-Cov2-infection.

|  | Not infected<br>N% | Infected<br>N% |
| --- | --- | --- |
| **Total number** | 218 | 186 |
| **Female** | 114 (52%) | 89 (48%) |
| **Age** |  |  |
| <18 years | 31 (14%) | 15 (8%) |
| 18-24 years | 30 (14%) | 20 (11%) |
| 25-39 years | 81 (37%) | 43 (23%) |
| 40-64 years | 71 (33%) | 100 (54%) |
| 65+ years | 5 (2%) | 8 (4%) |
| **BMI (kg/m<sup>2</sup>) (Std Dev)** | 24.3 (5.12) | 26.2 (5.16) |
| **Participating house hold member (Std Dev)** | 2.1 (1.12) | 2.4 (1.16) |
| **Highest level of formal education** |  |  |
| None | 27 (13%) | 13 (7%) |
| lower secondary school | 27 (13%) | 23 (13%) |
| secondary school | 55 (26%) | 71 (39%) |
| higher education entrance qualification | 54 (25%) | 34 (18%) |
| (technical) university degree | 52 (24%) | 43 (23%) |
| **duration of attendance [h] (Std Dev)** | 4.7 (2.06) | 5.8 (1.85) |
| **Service Team** | 4 (2%) | 22 (12%) |
| **on stage during event** | 80 (37%) | 62 (34%) |
| **member of "Council of 11"** | 6 (3%) | 18 (10%) |
| **on stage during "Finale"** | 26 (12%) | 48 (26%) |
| **Behavior during break** |  |  |
| remaining seated | 73 (36%) | 85 (48%) |
| going outside | 114 (55%) | 72 (39%) |
| **alcohol consumption [drink] (Std Dev)** | 11.3 (7.76) | 12.2 (7.40) |
| **former smoker** | 34 (16%) | 45 (24%) |
| **active smoker ?10 ciagrettes a day)** | 54 (25%) | 23 (12%) |
| **at least one comorbidity** | 29 (13%) | 28 (15%) |
| **avg. distance to other participants [m] (Std Dev)** | 9.2 (1.68) | 9.1 (1.70) |
| **distance to air inlet [m] (Std Dev)** | 6.1 (3.22) | 6.0 (3.30) |
| **distance to air outlet [m] (Std Dev)** | 4.8 (2.94) | 5.1 (2.87) |

Overall, 186 out of 404 individuals tested seropositive for IgG- and 161 for IgA-antibodies (**suppl. figure 1**). To confirm seropositivity we performed a plaque reduction neutralization assay (**suppl. figure 2**) demonstrating neutralizing activity against SARS-CoV-2 of their respective antibody responses. Given the low specificity of the IgA assay, IgA seropositivity was not further considered**Error! Bookmark not defined**.. As we tested for seropositivity 51 days after the superspreading event, we additionally performed SARS-CoV-2 RT PCR analysis from pharyngeal swabs to exclude potential recent infections. Indeed, 19 participants tested positive in RT-PCR, and were therefore not considered in the study as there was no likelihood of infection during the superspreading event. Overall, we found that 46.0% (95% CI: [41.2%; 51.0%]) tested seropositive who attended the event, which was significantly higher than the overall estimated infection rate in the same community at large at that time. Indeed, officially 3.1% of the community were reported as positive cases at that time and we estimated the infection rate as 15.5% (95% CI:[12.3%; 19.00%])**Error! Bookmark not defined**. for the community. Taken together, an estimated 46% of participants became infected during a single superspreading event.

No association between sex and risk of infection was found ((OR: 1.01 [0.65; 1.58]) for women). On average infected individuals had a higher body mass index (26·2kg/m^2^ compared to 24.3kg/m^2^ for uninfected individuals). Infected participants were more likely to be clustered living in the same household (**table 1**). Having at least one comorbidity, including lung disease (42.3%), cardiovascular disease (53.3%), neurological disease (16.7%), cancer (58.3%) or diabetes (80%), did not increase the risk of infection (OR: 0.64 [0.33; 1.26]). In conclusion, sex and comorbidities did not seem to affect the risk of infection. We next assessed whether age influenced the risk of infection at the event, considering sex, duration of attendance and common household as covariates. Comparison across age-categories showed a lower risk for children (OR: 0.31 [0.14; 0.69]), and also for young adults (18-25 years, OR: 0.53 [0.26; 1.09]) as well as adults between 25 and 40 years (OR: 0.48 [0.28; 0.85]) in comparison to older adults (40 to 65 years) (OR: 1, reference), while seniors had a slightly higher risk (older than 65 years, OR: 1.1 [0.31; 3.97]) (**figure 3**). Our data suggest that an additional 10 years of age are on average associated with 28% increased risk of infection (OR: 1.28 [1.10; 1.48]).

**Fig. 3.**
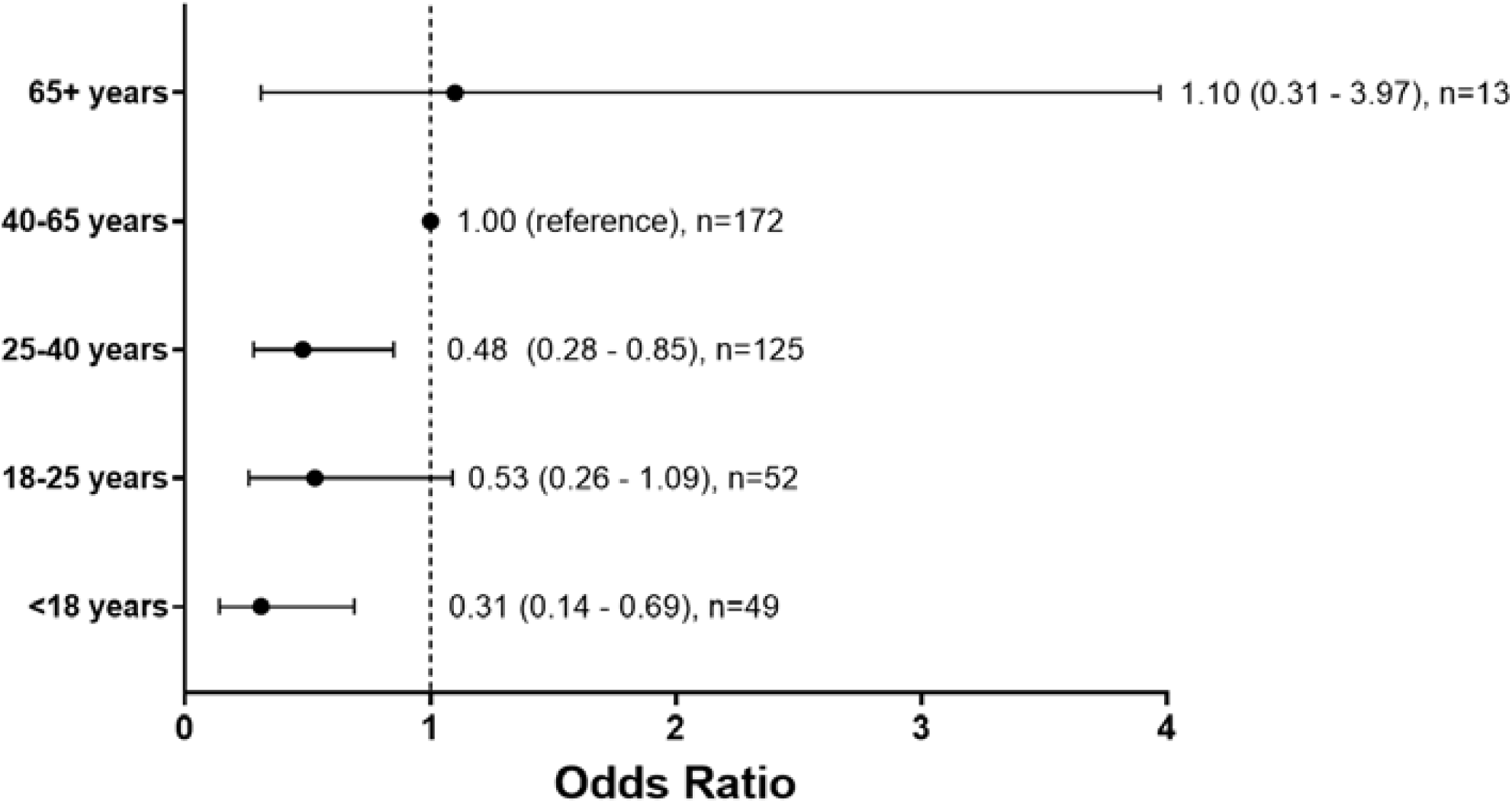
Odds-Ratio for the likelihood of SARS-CoV-2 infection by age groups. Participants were divided into age groups of 8, 15, or 25 years, participants younger than 18 or older than 65 years. Participants were considered to have been infected during the event if they were SARS-CoV-2 antibody positive (ELISA).

To understand the spreading dynamics of SARS-CoV-2 during the event, we first performed a detailed analysis of potential risk factors and social behavior. The event consisted of speeches, dance, and music performances for a total of five hours, with one large intermission and was hosted at a small community center (320 square meters) with a stage up front and a bar in the back close to the entrance. Alcoholic and nonalcoholic drinks were served in glasses and a food truck was located outside in front of the venue. While most participants were sitting in the hall, a committee of eleven individuals hosting the event were sitting on stage. The eleven people on stage switched after a break. With approximately 450 participants there were about 1·4 individuals per square meter and the tables, each with two benches, were arranged in two blocks with an alley to the stage (**figure 2**). Infected participants had been seated mostly at tables close to the bar, at the bar, or on stage. One table with 8 out of 11 infected people, was located far away from the bar at the other side of the hall and close to an air inlet. The group sitting on stage showed high numbers of infection (18 infected out of 24, **table 1**). We first analyzed whether the ventilation system influenced the distribution of SARS-CoV-2 infected individuals. It is important to state that the system’s air flow consisted of 75% used and 25% fresh air. The air flow can be described as clockwise. The air system uses vents along one side of the venue and on stage to take in air (**figure 2**, air inlets purple). After 25% of fresh air has been added and the air has been filtered, vents along the other side of the venue return the air into the room (**figure 2**, air outlets blue). All ventilation points received the same amount of air due to throttle valves. For noise protection reasons windows remained closed. The air-system used F7-Filters (ISO ePM ≥ 2,5) and had an air volume flow of 7500 m^3^/h.

Most tables located close to the air-inlets and showed no or only few infections (**figure 2**, green) also most surrounding tables showed low numbers of infection (**figure 2**, yellow). Tables close to the air-outlets (**figure 2)** show high (4 or 5 infected per table) and very high (6 or 7 infected per table) numbers of infected individuals. It is important to mention that the overall number of participants per table was not equal for all tables. Greater proximity to air outlets was associated with increased risk of infection with a crude OR=1.39 [0.86; 2.25]. This association remained stable and was hardly attenuated from adjustment for proximity to air inlet, age, gender, duration of attendance, proximity to other infected persons, stage-activity and going outside during the intermission (**figure 4**, multiple adjusted OR=1.26 [0.63; 2.50]). A similar apparent effect for proximity to air inlets (crude OR=1.17 [0.72; 1.89]) disappeared when duration of attendance was added to the model (**figure 4**, multiple adjusted OR=1.01 [0.53; 1.94]). Overall, however, we found the increased risk for individuals located closer to the air outlet remarkably persistent (**figure 4**).

**Fig. 4:**
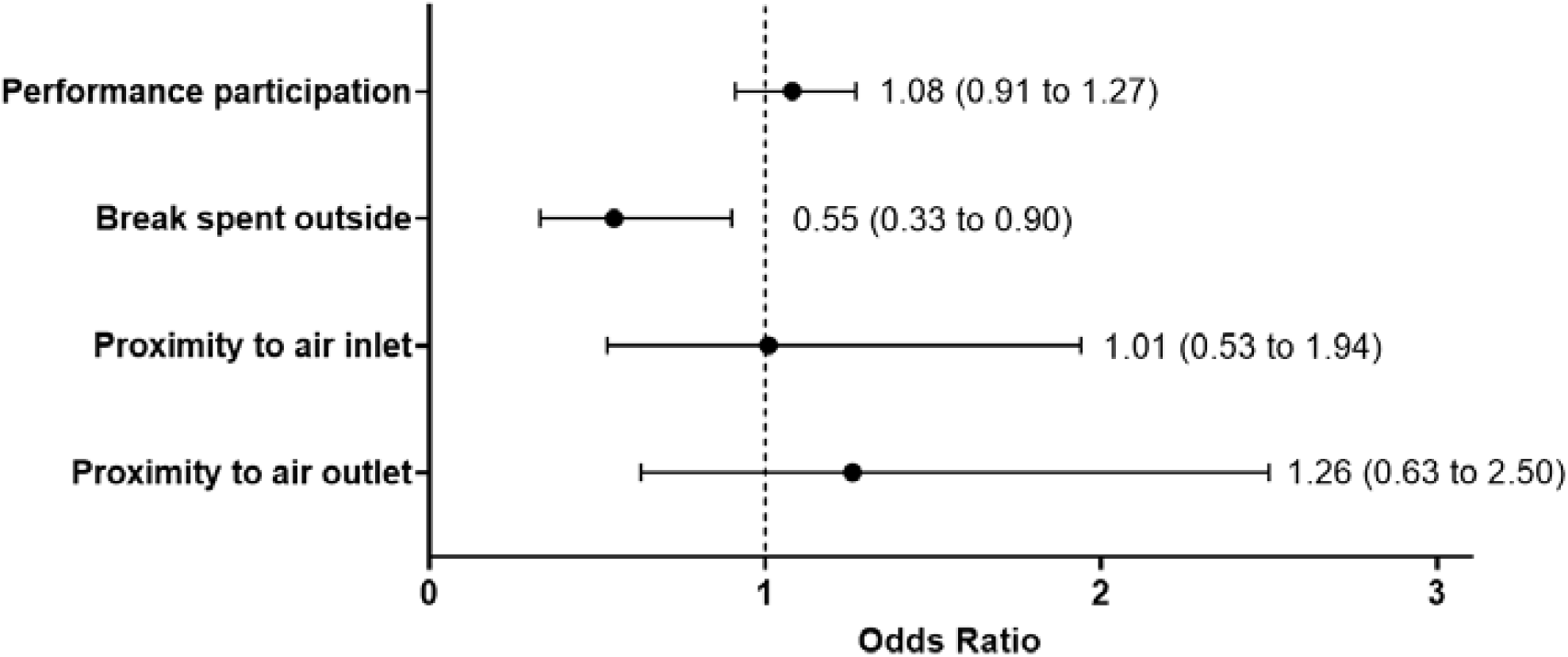
Odds ratios for the association of SARS-CoV-2 infection with specific activities of the participants and their location in the venue relative to ventilation shafts. The model was additionally adjusted for age, sex, duration of attendance, participation in multiple activities, and cumulative proximity to other infected persons, and common household.

We further studied the sum of the inverse distance to all infected participants as a measure of proximity to either one common virus source or mutual infection. However, there was no evidence for increased risk of infection from greater proximity to other infected persons (**suppl. table 1**). Furthermore, we found no evidence for a single person being the source of the infection using 401 quantile-plot analysis conducted for each participant as potential source of infection separately (**suppl. figure 3**).

To understand the association of risk with behavior patterns we next investigated the influence of several factors on SARS-CoV-2 infection including time spent outside, smoking, performing on stage and participation during the final act (“Finale”) for 30 minutes. Results were all adjusted for age, sex, common household, and duration of attendance. Participation in multiple performances did not increase the risk of infection (OR per performance: 1.08 [0.91; 1.27]) while participation in the last “Finale” indicated a trend towards increased risk of infection (OR: 1.41 [0.65; 3.02]) (**figure 4**). Duration of attendance was persistently and strongly associated with an increased infection risk of 32% with each additional hour spent at the party (OR per hour: 1.32 [1.16; 1.49]). All further analyses were adjusted for this variable as potential confounding factor.

We next determined the level of alcohol consumption as number of drinks (high-proof liquor or beer) and did not observe any influence for the amount of alcohol consumption on the risk of becoming infected (OR per drink: 1.00 [0.96; 1.05]). Furthermore, participants who spent the break outside were less likely to be infected (OR: 0.55 [0.33; 0.91]) compared to individuals who spent the break inside the venue hall (**figure 4**). Interestingly, however, when we determined the impact of being regular smoker (defined as smoking of at least 10 cigarettes a day) on the risk of SARS-CoV-2 infection we observed a reduced risk of infection (OR: 0.32 [0.12; 0.81]) even after adjustment for “time spent outside”. In conclusion, duration of attendance at the carnival party increased the risk of infection, the number of alcoholic drinks was not associated with infection risk, while regular smoking and spending the break of the event outside lowers the risk of infection.

We next stratified seropositive individuals by their reported symptoms. Odds-ratios for each symptom were calculated for the timespan of 14 days following the event (**figure 5**). Similar to previous reports^8^ loss of smell (OR: 8.78 [4.81; 16.02]) and taste (OR: 10.09 [5.13; 19.88]) were strongest associated with SARS-CoV-2 infection. Other symptoms which were strongly associated with COVID-19 were: sweats and chills (OR: 5.28 [3.08; 9.07]), muscle and joint ache (OR: 5.19 [3.19; 8.44]), fatigue (OR: 4.22 [2.76; 6.45]) and fever (OR: 3.73 [2.10; 6.63]) (**figure 5**). Importantly, 15.1% of the infected individuals reported no symptoms at all in a period of 14 days after the event. The rate of asymptomatic infections of participants of the event was lower than generally observed in the community the event took place in (36%).^5^ Overall, there was a lower proportion of asymptomatic cases among individuals infected after the event compared to members of the community, while loss of smell and taste showed the strongest association with an infection.

**Fig. 5:**
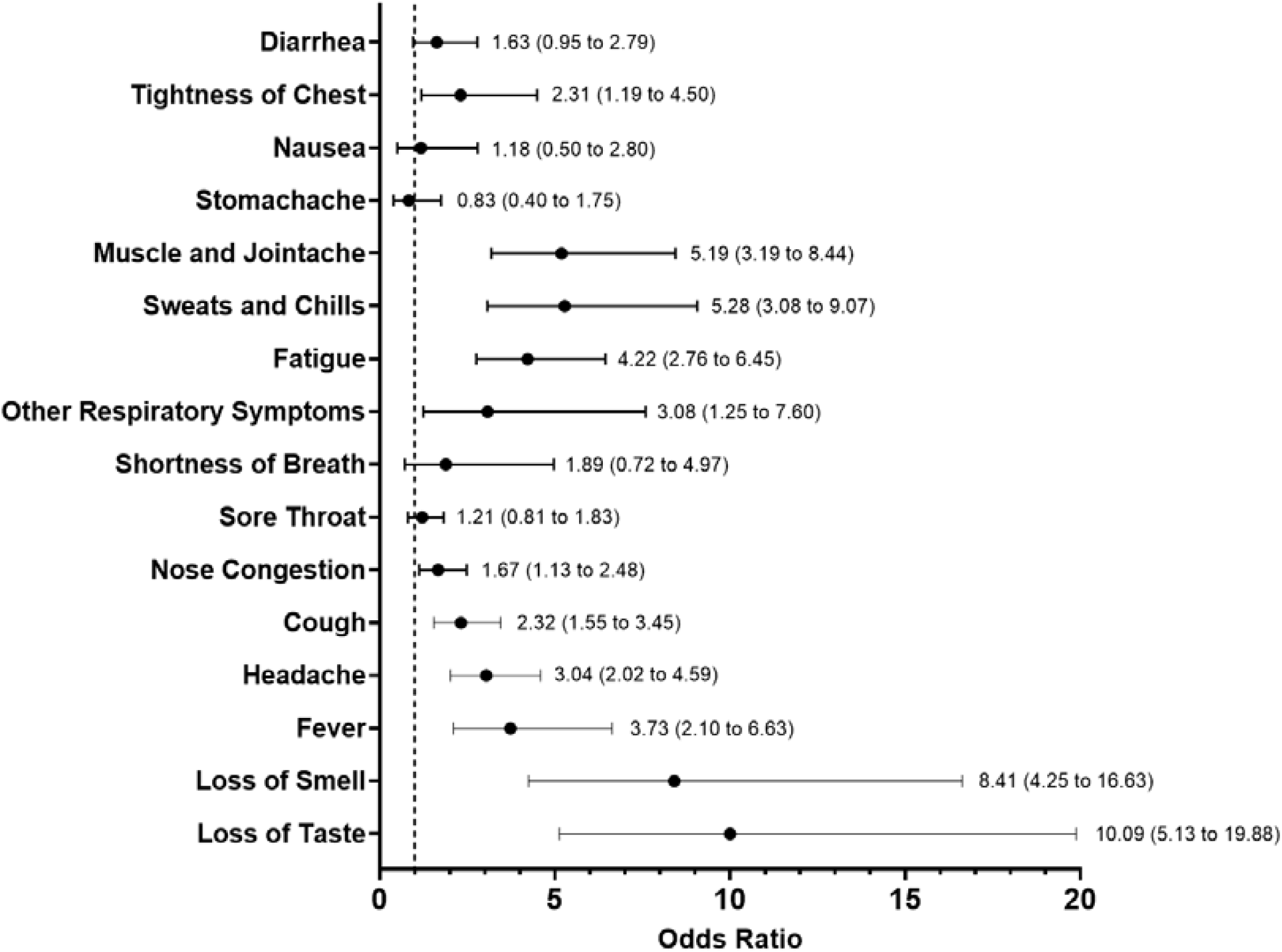
Odds ratios for symptoms of SARS-CoV-2 antibody-positive participants in the 14 days following the super spreading event. The information on symptoms was derived from the self-administered questionnaire, which was filled out on the day of sample collection. Odds ratio estimates (OR) are shown with confidence intervals

## Discussion

The high overdispersion characteristics of SARS-CoV2 and its ability to be transmitted via aerosols under certain conditions are one of the main reasons that the beginning of the SARS-CoV2 pandemic was shaped by superspreading events.^9,10^ Germany’s first superspreading event was an indoor carnival event in the beginning of 2020 in a rural the community. In this naturally occurring experiment, we demonstrate that nearly half of the participants became infected and demonstrate multiple prerequisites of such an event and risk factors for becoming infected. While our study population is not a representative sample of the general population the event may be regarded as exemplary for similar party occasions and may help reduce the number of infected in the future.

An important factor associated with infection risk was the ventilation system and the individual proximity to the ventilation outlets. Individuals close to the air-outlets that contained air with low amount of fresh air had the highest infection risk compared to those close to the air-inlets. This is in line with previous studies that demonstrated SARS-CoV-2 to be able to become air-borne under certain conditions and that the ventilation system can have an influence on virus spread.^11,12,13^ The air filters in the venue were not capable of intercepting virus particles supporting the notion on the importance of proper indoor ventilation systems.^14,15^ Indeed, spending the break of the event outside decreased the possibility of infection underscoring the benefit of proper ventilation to lower the amount of aerosols. Due to the nature of the event, the spatial distribution of the participants was not fixed throughout the evening, and not perfectly recapitulated, so this information carries some error. However, allowing for multiple positions per person we used all available information. Assuming further error in the spatial data to be random, this might lead to a dilution of effects, i.e. true associations may remain undetected. Complementary analyses including e.g. the persons’ functions during the event show consistent results, so we see no evidence suggesting bias in our findings.

The consumption of alcoholic drinks did not increase the risk of infection. While it has been assumed that the alcoholic effect of decreased social inhibition may increase likelihood of infection, we did not find any evidence for this association questioning measures of a ban on alcohol to reduce numbers of infected. It is known that current and former smokers disproportionately suffer from severe COVID-19 and their numbers are relatively increased among those patients that need intensive care treatment compared to non-smokers.^16,17^ However, it has been previously speculated that the risk of infection is lower for smokers.^18^ Furthermore, a meta-analysis of seven studies suggests that smokers have a reduced risk of testing positive for SARS-CoV-2.^19^ Interestingly, we also observed a protective effect for an infection with SARS-CoV-2, thus our findings support those statements and show an even greater protective effect. The association might for example be explained by a role of the nicotinic acetylcholine receptor.^20^ While we strongly advise that smoking should not be considered as a protective habit to prevent risk of infection, this knowledge may lead to the investigation of a therapeutic or prophylactic treatment on the basis of this molecular target.^21^

Our results indicate a trend that younger people are less likely to be infected compared to older age groups. This trend is strongest for people under 18 but levels out over 40 years of age. The risk of infection for children in superspreading events has not been investigated but the overall risk for infection in children seems to be lower than for adults as a systematic review and its recent update reported, which is further supported by our findings.^22,23^ As all individuals were exposed at the same event and time our study is a perfect model for the previously described notion, that children are less likely to become infected. Indeed, a recently published meta-analysis by Viner et al. showed a low susceptibility for children and adolescents (OR of 0.56 (95% CI, 0.37-0.85)) which strongly supports our findings of a lower risk of infection in that age group, which is even lower in our study.^24^ Our finding supports the previously shown subordinate influence on the spreading of the virus by children. The finding that each 10 years of age increase the risk of infection during an event indicates that younger people and their limited role should be considered when measures to contain the pandemic are implemented. Taken together, we could demonstrate important risk factors for infection during a superspreading event, which helps to understand transmission dynamics in order to improve comprehensive public health preparedness measures.

## Data Availability

The data contain information that could compromise the privacy of research participants. Data sharing restrictions imposed by national and trans-national data protection laws prohibit general sharing of data. However, upon submission of a proposal to the principal investigator of this study and approval of this proposal by (i) the principal investigator, (ii) the Ethics Committee of the University of Bonn and (iii) the data protection officer of the University Hospital Bonn, data collected for the study can be made available to other researchers. The Insitute of Virology, University Hospital Bonn has full access to all of the data.

## Acknowledgments

We would like to thank the inhabitants of the town in question for their participation. We would also like to thank the local government of the town where the outbreak took place for their support to conduct the study. Furthermore, we would like to thank the following people who helped with the study: Rudi Kirschen, Gero Wilbring, Janett Wieseler, Marek Korencak, Ryan Nattrass, Jernej Pusnik, Maximilian Becker, Ann-Sophie Boucher, Marc Alexander de Boer, Rebekka Dix, Sara Dohmen, Kim Friele, Benedikt Gansen, Jannik Geier, Marie Gronemeyer, Sarah Hundertmark, Nora Jansen, Michael Jost, Louisa Khorsandian, Simon Krzycki, Ekaterina Kuskova, Judith Langen, Silvia Letmathe, Ann-Kathrin Lippe, Jonathan Meinke, Freya Merker, Annika Modemann, Janine Petras, Sophie Marie Porath, Anna Quast, Laurine Reese, Isabel Maria Rehbach, Jonas Richter, Thea Rödig, Eva Schmitz, Tobias Schremmer, Louisa Sommer, Jennifer Speda, Yuhe Tang, Oliver Thanscheidt, Franz Thiele, Johanna Thiele, Julia Tholen, Moritz Transier, Maike van der Hoek, Tillmann Verbeek, Sophia Verspohl, Kira Vordermark, Julian Wirtz, Marina Wirtz, Lisa Zimmer, Philip Koenemann, Adi Yaser, Lisa Anna, Katharina Bartenschlager, Lisa Baum, Roxana Böhmer Romero, Diana de Braganca, Isabelle Engels, Moritz Färber, Carina Fernandez Gonzalez, Lucia Maria Goßner, Victoria Handschuch, Franziska Georgia Liermann, Steffan Meißner, Laura Racenski, Patrick, Denis Raguse, Larissa Reiß, Maximilian Rölle, Franziska Scheele, Chiara Schwippert, Arlene Christin Schwippert, Antonia Seifert, Joshua David Stockhausen, Sofia Waldorf, Leonie Weinhold, Nicolai Trimpop, Julia Reinhardt, Vera Gast, Michelle Yong, Eva Engels, Jonathan Meinke, Susanne Schmidt, Janine Schulte, Saskia Schmitz, Kübra Bayrak, Regina Frizler, Katarzyna Andryka, Sofía Soler, Bastian Putschli, Thomas Zillinger, Marcel Renn, Patrick Müller, Dillon Corvino, Zeinab Abdullah, Katrin Paeschke, Hiroki Kato, Florian Schmidt, Maximilian Baum, Celina Schüter, Daniel Hinze, Martina Schmidt, Arcangelo Ricchiuto, Sonja Gross, Uta Wolber, Tobias Höller, Marion Zerlett, Esther Sib, Benjamin Marx, Souhaib Aldabbagh. We thank the Medizinischer Dienst der Krankenkasse (MDK) Nordrhein for their generous help in conducting this study. In particular we would like to thank: Tanja Bell, Ina de Hesselle-Taddey, Susanne Goll, Katja Hoffmann-Pruss, Niko Kalamakidis, Verena Kayser, Hildegard Kessler, Norbert Körfer, Daniela Kroll, Florian Messerschmidt, Svenja Peters, Sebastian Schröder, Linda Schroers, Michael Zimmermann, Klaus-Peter Thiele. We thank Stefan Holdenrieder, Alexander Semaan, Bernd Pötzsch, and Georg Nickenig for providing control samples. The idea, the plan, the concept, protocol, the conduct, the data analysis, and the writing of the manuscript of this study were independent of any third parties, including the government of North Rhine-Westphalia, Germany.

